# COVID-19 testing: disparity between national and institution-based case detection

**DOI:** 10.1101/2022.02.21.22270847

**Authors:** Timothy J Aitman, Linda Bauld, Kathryn F Carruthers, Nick Gilbert, Neil Turok

**Affiliations:** Centre for Genomic and Experimental Medicine, Institute of Genetics and Cancer, University of Edinburgh, Edinburgh, EH4 2XU, UK; NHS Lothian, Edinburgh, EH4 2XU, UK; Usher Institute, College of Medicine and Veterinary Medicine, University of Edinburgh, EH8 9AG, UK; School of Physics and Astronomy, College of Science and Engineering, University of Edinburgh, EH9 3FD, UK; MRC Human Genetics Unit, Institute of Genetics and Cancer, University of Edinburgh, EH4 2XU, UK; Higgs Centre for Theoretical Physics, James Clerk Maxwell Building, University of Edinburgh EH9 3FD, UK

## Abstract

Reports of COVID-19 prevalence through national statistics, community surveys and targeted testing at places of work or study have guided national and institutional responses to the pandemic. The University of Edinburgh established a mass testing programme, TestEd, for detection of COVID-19 in asymptomatic staff and students who are studying or working on campus. The study has tested more than 100,000 samples with more than 170 confirmed positive results. Since the introduction of a change in policy in England and the UK devolved nations in early January 2022, to limit eligibility for PCR testing in the community to those with symptoms, we have noticed a divergence between the reports in Scottish and UK-wide prevalence, and the magnitude and frequency of positive results in the University datasets. While the national UK-wide and Scottish case figures show declining or stable prevalence, University case reports have risen more than five-fold since early December 2021 and continue to rise. These observations could be important in the face of future variants of concern and emphasise the need for continued access to high sensitivity PCR testing and other forms of surveillance.

## Introduction

Reports of COVID-19 prevalence through national statistics, community surveys and targeted testing at places of work or study have guided national and institutional responses to the pandemic. However, strategies for infection control targeted solely at individuals with symptoms are unlikely to be completely effective at containing viral spread, because around 50% of transmission has been observed to occur from pre-or asymptomatic individuals.^1^

The University of Edinburgh established a mass testing programme, TestEd,^2^ for detection of COVID-19 in asymptomatic staff and students who are studying or working on campus. Because TestEd participants don’t have symptoms, individuals in this population who are pre-or asymptomatic carriers would not realise the risk they pose in transmitting the virus to colleagues or peers in the absence of a positive test. In this report, we compared the frequency of positive test results from the TestEd programme, from March 2021 - February 2022, with the reports of positive cases from the University of Edinburgh reporting system and published national data from Public Health Scotland.

## Methods

Data was collected from three different sources.

i. The University of Edinburgh mass testing programme, TestEd, uses pooled saliva-based testing by PCR, with a protocol adapted from our previously published approach for nasopharyngeal swab testing.^3^ The study has tested more than 100,000 samples with more than 170 confirmed positive results. Specificity is > 99% and sensitivity, based on controlled samples, is 93-97%. Results are reported daily but collated and presented weekly.
ii. UoE cases: The University collates data from across the University according to University guidelines requiring anyone testing positive via any route (LFD, NHS test, Tested) to report this on a standard online form to the University. This data was collated on a weekly basis and tests from all routes were combined to give a weekly report of positive cases within the University. A small proportion of positive cases reported to the University came from TestEd. However, because the TestEd protocol aimed routinely to test only asymptomatic participants and the majority of cases reported to the University were largely symptomatic, the overlap is small (less than 5% of overall University cases) and positive cases that were present in both datasets were not removed.
iii. Daily reports of positive cases were downloaded from the COVID-19 Daily Dashboard on the Public Health Scotland web site and collated to give a weekly case report.

## Results

Since the introduction of a change in policy in England and the UK devolved nations to limit eligibility for PCR testing in the community to those with symptoms, we noticed a divergence between the reports in Scottish and UK-wide prevalence, and the magnitude and frequency of positive results in the University datasets. This change in eligibility for PCR tests took effect in Scotland from 6^th^ January 2022, following which national data reporting was adjusted from 13^th^ January 2022 to reflect this revision.^4^ Cases are now a combination of self-reported lateral flow tests (intended for asymptomatic individuals in the community) and those with symptoms who are still encouraged to seek a PCR test.

The national [UK-wide](https://www.ons.gov.uk/peoplepopulationandcommunity/healthandsocialcare/conditionsanddiseases/bulletins/coronaviruscovid19infectionsurveypilot/4february2022) and [Scottish](https://publichealthscotland.scot/publications/covid-19-statistical-report/covid-19-statistical-report-2-february-2022/) case figures show declining or stable prevalence. In contrast, University case reports have risen more than five-fold since early December 2021 and continue to rise (Figure). A similar rise was observed in both University staff and students. The TestEd result, from a relatively stable sample of regular, asymptomatic participants, may be less prone to reporting biases than the present national statistics.

**Figure.**
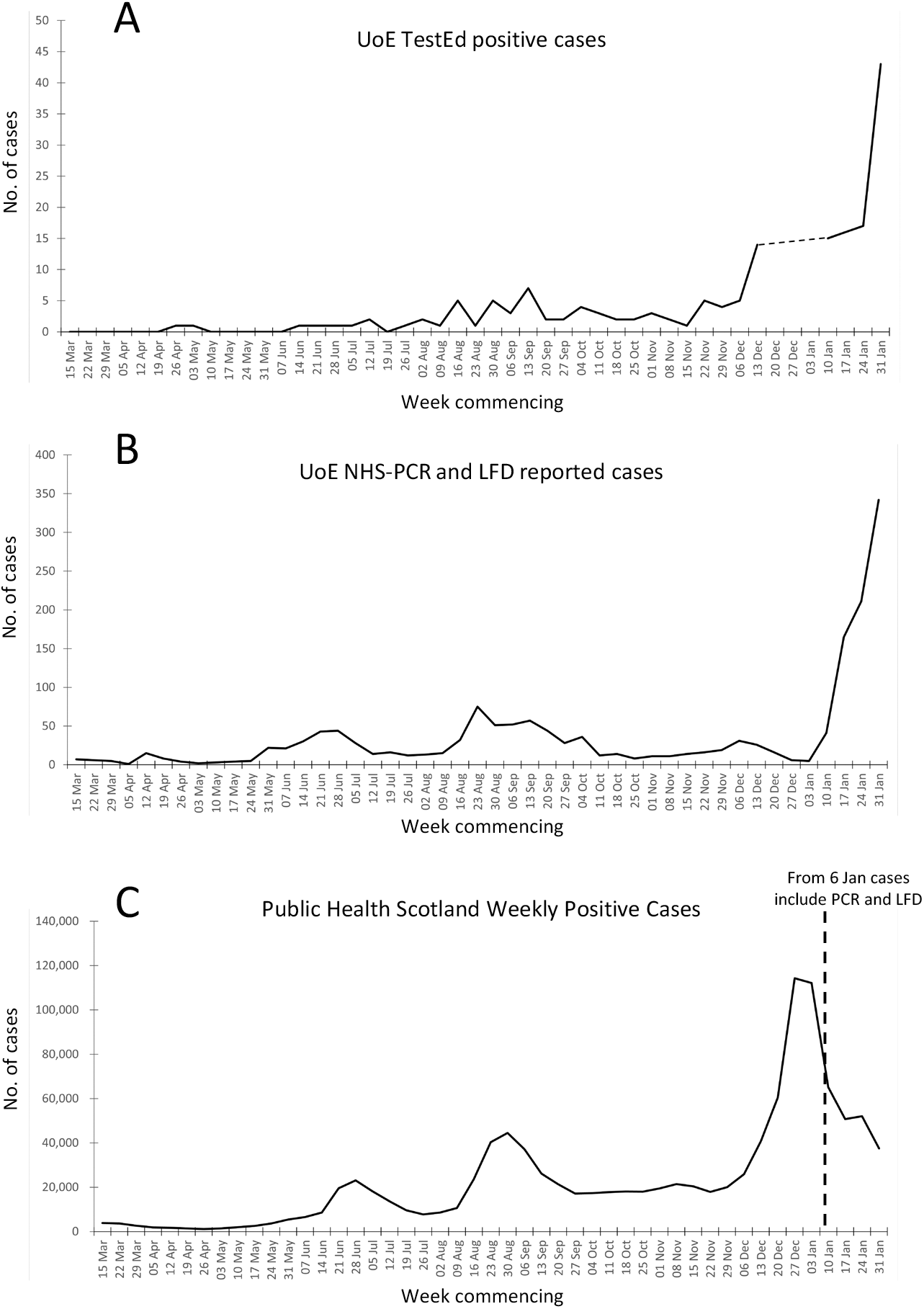
Reported COVID-19 cases, 15th March 2021 - 4th February 2022, from (A) University of Edinburgh TestEd programme, (B) University of Edinburgh total positive NHS and LFD tests, (C) Public Health Scotland reports. The discontinuity in TestEd data (dashed line) is due to Christmas closure of the TestEd laboratory.

## Discussion and Conclusions

Our PCR and sequencing data and analysis of the demographics of the individuals testing positive suggest that the rise in prevalence in the University population is reflective of a substantial increase in viral prevalence that is not reflected in nationally reported positive case numbers. This has also been observed in other UK-wide data sources including the ZOE app study.^5^ Although there are potential sources of data bias, including increased testing amongst mildly symptomatic cases and local changes in viral strain, we suggest these results are most likely due to reduced testing and, in particular, a dearth of reporting of positive LFT results amongst the COVID-positive population. While hospital admissions from COVID-19 are declining due to high rates of vaccination, protection from prior infections and a reduced severity of the Omicron variant, public health messaging needs to acknowledge that reduced eligibility for PCR testing and under-reporting of positive LFTs can lead to under-estimates of community prevalence. This could be important in the face of future variants of concern, emphasising the need for continued access to high sensitivity PCR testing and other forms of surveillance. These should prioritise occupational or high risk settings and vulnerable populations, along with population surveillance via wastewater testing and community-based studies.

## Data Availability

All data produced in the present work are contained in the manuscript.

## Authors’ contributions

TJA, LB, NG and NT conceptualised and designed the study. TJA and KFC collated and analysed the data. TJA, LB and NG wrote the manuscript. All authors reviewed and approved the manuscript.

## Conflict of interest statement

Declarations of interest: LB is interim chief social policy adviser to the Scottish government. TA is Founder and Director of the company BioCaptiva Ltd. All other authors declare no interests.

## Role of funding source

TestEd is funded by UKRI Research Grant MR/W006243/1 and the University of Edinburgh.

## Ethics committee approval

The study was carried out under Edinburgh Medical School Research Ethics Committee approved protocol Ref 20-EMREC-023_SA04.

## Notes

### Competing Interest Statement

Mixed competing interests. All authors have completed the ICMJE uniform disclosure form at www.icmje.org/coi_disclosure.pdf and declare: TA undertakes consultancy for BioCaptiva Ltd (unrelated to present manuscript). TA received support for attending meetings and/or travel from Global Engage and Oxford Global. TA has a patent filed in the liquid biopsy field unrelated to the present study. TA participates in the Scottish Genomics Leadership Group Advisory Board. TA was a panel member of the European Research Council (2018 to 21). TA is a shareholder of BioCaptiva Ltd (unrelated to present work).
LB serves as interim chief social policy adviser to the Scottish Government and is a member of the Chief Medical Officers (Scotland) Advisory Group on Covid19.

### Funding Statement

The study was funded by the University of Edinburgh and by UKRI Research Grant MR/W006243/1.

### Author Declarations

Edinburgh Medical School Research Ethics Committee of University of Edinburgh gave ethical approval for this work (EMERC21).

